# Seroincidence of typhoidal *Salmonella* infection in eastern Democratic Republic of the Congo: a population-based cross-sectional serosurvey

**DOI:** 10.64898/2026.11.25.26368987

**Authors:** Jonathan Polonsky, Lise Grout, Mahamadou Maiga, Benard Gacheru, Gabriel Alcoba, Patrick Nkemenang, Octavie Lunguya, Andrew S. Azman, Primitive Gakima, Andrea Incerti, Etienne Gignoux, Kristen Aiemjoy, Jessica C. Seidman, Iza Ciglenecki, Richelle C. Charles

## Abstract

**Background:** The Democratic Republic of the Congo (DRC) lacks population-based estimates of the incidence of typhoidal *Salmonella* infection, despite clinical and mortality evidence of a high burden, and has yet to introduce typhoid conjugate vaccine (TCV). We estimated seroincidence in conflict-affected Angumu Health Zone, Ituri Province.

**Methods:** We conducted a cross-sectional serosurvey across 14 health areas within an operational malaria intervention survey in April–May 2024. Seroincidence was estimated from IgA responses to haemolysin E (HlyE) in dried blood spots (DBS) with models of post-infection antibody decay. Seropositivity was the proportion of participants with HlyE IgA at or above a cut-off indicating recent infection. Estimates were weighted by intervention-zone population.

**Findings:** We included 2586 participants from 582 households. Overall IgA seroincidence was 25 infections per 1000 person-years (95% CI 22–29), peaking in children younger than 2 years (91, 95% CI 46–180) and exceeding 20 per 1000 person-years in every older age group. The differences in age-standardised seroincidence by sex (28 in male and 22 in female participants, p=0.051) and by self-reported typhoid history among participants aged 5 years or older (p>0.99) were not statistically significant. HlyE IgA seropositivity was 3.1% (95% CI 2.4–4.0), with smoothed seropositivity ranging from 2.4% to 7.2% across the study area.

**Interpretation:** These findings indicate substantial typhoidal *Salmonella* transmission in Angumu that clinical surveillance alone could not detect. Among children, seroincidence was higher than in serosurveys in Nepal but lower than in Bangladesh and Pakistan, and among adults it was lower than in all three. Because seroincidence includes mild and asymptomatic infections, it cannot be mapped directly onto WHO clinical-incidence thresholds for TCV introduction. Integrating DBS serosurveillance into an existing household survey was operationally feasible. These findings support consideration of targeted TCV introduction in similarly under-surveilled, high-transmission settings in eastern DRC.

**Funding:** Médecins Sans Frontières, Operational Centre Geneva.

**Research in context:** *Evidence before this study:* We searched PubMed and Embase from database inception to Nov 25, 2026, without language restriction, for population-based estimates of typhoidal Salmonella infection in central Africa. We combined terms for typhoid and enteric fever with terms for seroincidence, serosurvey, and haemolysin E, and with terms for the Democratic Republic of the Congo and for Africa. Incidence estimates for the DRC come from facility-based surveillance, which counts the patients who reach a hospital with a functioning blood-culture laboratory. Seroincidence estimated from antibody responses to haemolysin E measures infection rather than clinical disease, and has been reported from Juba in South Sudan, from coastal and western Kenya, and from cohorts in south Asia. The searches returned no seroincidence estimate for the DRC, and none from a conflict-affected population in central Africa.

*Added value of this study:* This is the first population-based estimate of typhoidal Salmonella seroincidence in the DRC. Infection was frequent in every age group, and highest in children younger than 2 years, in a health zone where no typhoid case has been confirmed by culture. Collecting dried blood spots within an existing operational household survey, and analysing them with published antibody-decay models, produced these estimates without dedicated surveillance infrastructure.

*Implications of all the available evidence:* Typhoidal Salmonella transmission can be intense where no diagnostic capacity exists to record it, so the absence of confirmed cases is not evidence of a low burden. Serosurveillance embedded in household surveys can supply the local burden data that countries need in order to decide where to introduce typhoid conjugate vaccine. National immunisation programmes and their partners could use it for that purpose in eastern DRC.

## Introduction

Enteric fever, a systemic infection caused by *Salmonella enterica* serovars Typhi and Paratyphi, causes an estimated 9.3 million infections and over 100,000 deaths annually,^1^ with the highest burden borne by populations in low- and middle-income countries with inadequate water, sanitation, and hygiene (WASH) infrastructure.^2^ Children are disproportionately affected, and the burden among those younger than 5 years may be higher than previously recognised.^1,3,4^

Antimicrobial resistance in *S*. Typhi increasingly limits treatment options, which strengthens the case for prevention through vaccination.^5,6^ Licensed vaccines target *S*. Typhi specifically; no vaccine is currently available for *S*. Paratyphi. Typhoid conjugate vaccine (TCV), which can be given from 6 months of age, showed 78–85% efficacy against blood culture-confirmed typhoid in phase 3 trials among children in Nepal, Bangladesh, and Malawi.^7–10^ The 2026 World Health Organization (WHO) position paper recommends TCV where typhoid incidence is high or very high, the case fatality ratio is high, or antimicrobial resistance is common.^11^ Gavi has supported TCV introduction in several countries.^12–14^

Nevertheless, uptake has been uneven, and large parts of sub-Saharan Africa are without access to TCV.^14^ The Democratic Republic of the Congo (DRC), a Gavi-eligible country, has not introduced TCV into its national immunisation programme.^15^ In the Severe Typhoid in Africa (SETA) programme, facility-based clinical surveillance at two health centres in Kisantu yielded an adjusted incidence of 3.2 symptomatic cases per 1000 person-years, the highest of the six countries.^6^ SETA investigators also documented antimicrobial resistance and intestinal perforation among hospitalised patients in the DRC.^6^ No community-based estimate of the incidence of typhoidal *Salmonella* infection is available for the DRC. A lack of population-based burden data has been a barrier to TCV introduction decisions.^12^

Blood culture, the gold standard for typhoid diagnosis, is rarely available in resource-constrained settings and detects approximately 60% of true cases.^16^ Serological approaches offer an alternative. Seroincidence, the rate of new *S*. Typhi and *S*. Paratyphi A infections per person-time, can be estimated from cross-sectional, population-based samples.^4^ The estimate is inferred from the distribution of antibody levels in the sample together with models of the rise and decay of haemolysin E (HlyE) antibodies after infection. Seropositivity is the proportion of participants whose HlyE IgA level is at or above a cut-off indicating recent infection. Because HlyE is expressed by both *S*. Typhi and *S*. Paratyphi A, these estimates cannot distinguish infections caused by the two pathogens.

Angumu Health Zone, in Ituri Province in north-eastern DRC, has experienced recurrent conflict and displacement since 2017.^17^ Clinicians at the hospital serving the health zone, where surgical capacity is limited, have observed many patients with intestinal perforation consistent with severe enteric fever. At that hospital in 2023, peritonitis accounted for 18% of the approximately 45 operations each month, and 6 of the 8 monthly cases were in children (MSF, unpublished data). Typhoid and paratyphoid fever accounted for 4.2% (95% CI 3.1–6.2) of deaths among people aged 12 years or older in a 2022 verbal autopsy study in the same population.^18^ Enteric fever was among the ten leading causes of adult death assigned by clinician review. The DRC meets WHO criteria for priority introduction of TCV.^15^

In Angumu Health Zone, between April and May 2024, we estimated the seroincidence of typhoidal *Salmonella* infection as a baseline before any vaccination campaign. The primary outcome was HlyE IgA measured in dried blood spots. As secondary analyses, we estimated seroincidence from combined HlyE IgA and IgG and described spatial variation in HlyE IgA seropositivity.

## Methods

### Study design and setting

From April to May 2024, we conducted a cross-sectional population-based serosurvey in Angumu Health Zone, Ituri Province, DRC. This serosurvey was embedded in a household survey evaluating the effect of malaria prevention interventions on mortality and malaria prevalence.^19^ On Lake Albert’s western shore, Angumu Health Zone comprises 22 health areas (Figure 1). Insecurity restricted sampling to 14 health areas east of the Kakoyi River (estimated population approximately 172,000).^17,20^ In 2022, these 14 areas were allocated to three malaria intervention zones serving as sampling strata. These zones were routine care (Routine; 5 health areas), indoor residual spraying (IRS; 4), and mass drug administration combined with IRS (MDA+IRS; 5). The study population comprised residents of selected households, including internally displaced persons.

**Figure 1.**
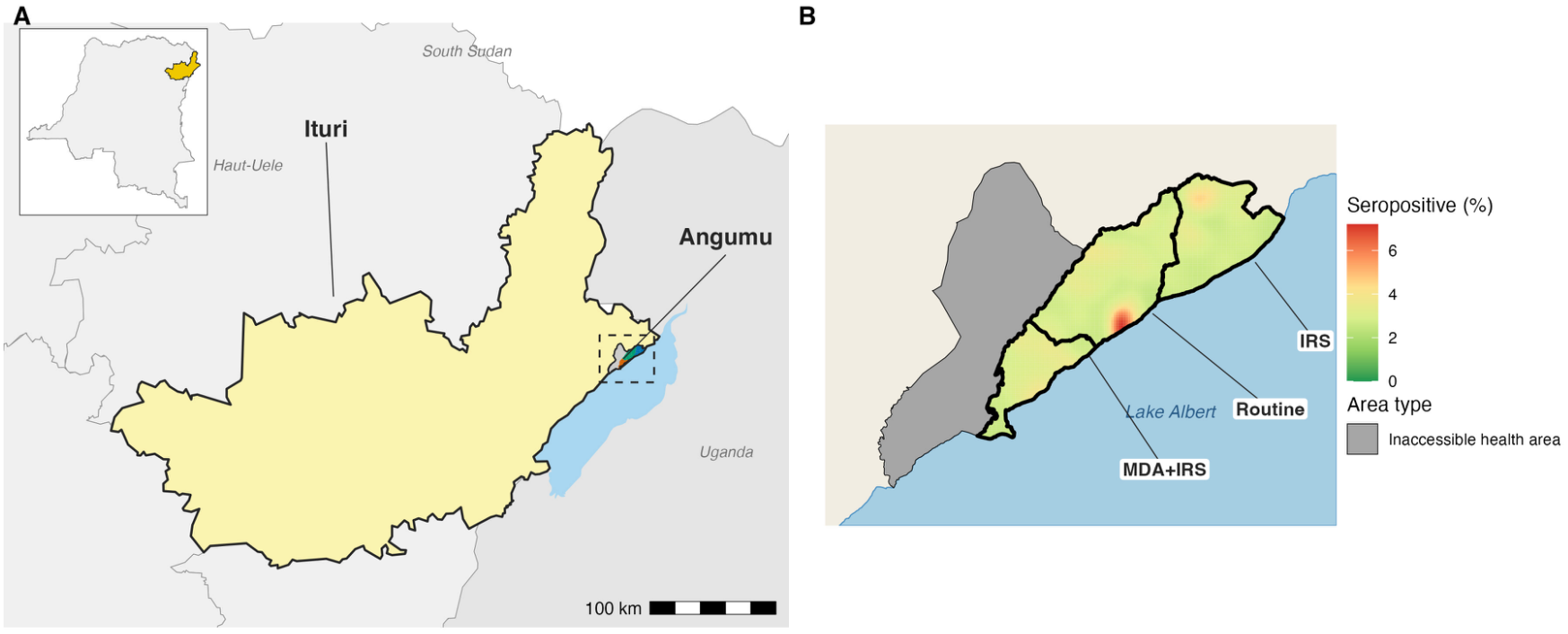
Study area and HlyE IgA seropositivity, Angumu Health Zone, Ituri Province, DRC. (A) Location of Angumu Health Zone within Ituri Province and the DRC. (B) Smoothed surface of HlyE IgA seropositivity across the sampled health areas, with the 2022 malaria intervention zones labelled. No household locations are shown.

### Participants and sampling

Within each zone, roofs were drawn at random from a satellite-image frame; each drawn roof and its four nearest neighbours formed a cluster of five households for the malaria survey, and one household per cluster joined the typhoid component (Supplementary Methods SM2). All household members who had lived in the household during the preceding 2 weeks were eligible, with no minimum age. Dried blood spot (DBS) samples were collected from consenting participants, or from children whose parent or legal guardian provided consent. Households with no member available after two visits, or in which participation was refused, were not replaced.

For the typhoid component, 3,062 individuals were required, assuming 3% recent-infection prevalence and ±20% relative precision for the overall estimate. The sample size was not calculated for age-specific estimates. To meet the malaria and serological requirements, 630 households (210 per zone) were selected.

### Data collection and laboratory procedures

Data were collected using standardised electronic questionnaires on tablets. Trained survey enumerators administered a rapid diagnostic test (RDT) for *Plasmodium falciparum* histidine-rich protein 2 (SD Bioline Ag P.f.; Standard Diagnostics, Republic of Korea).

Participants with a positive RDT were treated following national protocols. Consenting participants then provided a capillary blood sample from the same finger prick.

Approximately 60 µL of blood was collected on TropBio™ filter paper, air-dried for at least 30 minutes, and stored at −20 °C. Samples were shipped at room temperature to Massachusetts General Hospital, Boston, USA, and returned to −20 °C upon receipt until testing.

Between 7 May and 7 August 2025, DBS samples were eluted and tested for immunoglobulin A (IgA) and immunoglobulin G (IgG) responses to HlyE. We used kinetic enzyme-linked immunosorbent assays (ELISA) following published protocols.^4,21^ Elution, detection, and normalisation to ELISA units (EU) are described in Supplementary Methods SM1.

### Outcomes, variables, and definitions

The primary outcome was typhoidal *Salmonella* seroincidence estimated from HlyE IgA, based on expert consensus that HlyE IgA kinetics are more stable across settings with different forces of infection.^22^ Combined HlyE IgA and IgG estimates were secondary, and HlyE IgG-only estimates were exploratory. HlyE IgA seropositivity was defined as ≥9.23 normalised EU, a published diagnostic cut-off indicating recent infection.^23^ Age groups (<2, 2-4, 5-15, and ≥16 years; Supplementary Methods SM7) followed the clinical enteric fever surveillance bands of the published kinetics.^4,23^ The <2 and 2-4 year groups shared the <5 kinetics. History of typhoid fever was self-reported (question wording in Table 1), although self-reported history may not reflect laboratory-confirmed disease because diagnostic capacity is limited.

**Table 1.** Characteristics of participants included in the seroincidence analysis, Angumu Health Zone, April–May 2024.

| Characteristic | N = 2,586 |
|---|---|
| Total | 2586 |
| Age group |  |
| <2 | 130 (5.0%) |
| 2-4 | 371 (14%) |
| 5-15 | 979 (38%) |
| 16+ | 1,106 (43%) |
| Sex |  |
| Female | 1,393 (55%) |
| Male | 1,159 (45%) |
| Missing | 34 |
| Self-reported typhoid history |  |
| No | 2,093 (81%) |
| Yes | 488 (19%) |
| Missing | 5 |
| Household context |  |
| Households contributing a specimen | 582 |
| Participants sampled per household, mean (range) | 4.4 (1-16) |
| Household size at interview, mean (range), among 2504 participants with a matched household (564 households) | 5.2 (1-17) |

Self-reported typhoid history: answer to “Has [name] ever been ill with typhoid fever?” Participants answering yes were asked who had said it was typhoid fever; among the 472 with a linked survey record, this was a doctor or nurse for 365 (77%), a pharmacist for 57 (12%), and unknown for 41 (9%). The year of illness was also asked but not analysed.

### Statistical analysis

We estimated seroincidence by maximum likelihood, comparing observed quantitative HlyE IgA and IgG distributions with those expected under different infection rates.^4,24^ Age-specific antibody kinetics were pooled from longitudinal blood-culture-confirmed enteric fever cohorts in Bangladesh, Nepal, Pakistan, and Ghana. We used the *serocalculator* package (version 1.4.1) in R (version 4.5.2).^25^ We report seroincidence as infections per 1,000 person-years with 95% confidence intervals. Measurement-noise parameters were those estimated for the testing laboratory. Kinetic and noise parameters are given in Supplementary Table S1, and assumed antibody curves with Angumu titres in Supplementary Figure S1. Those kinetics came mostly from dried blood spots collected and dried as here, with plasma at enrolment.^4^ To assess sensitivity to these inputs, we refitted the unweighted IgA analysis with alternative kinetic and measurement-noise parameters.

We used the *serocalculator* cluster-robust variance estimator for within-household clustering. To assess household clustering, we repeated the unweighted IgA analysis with one randomly selected participant per household and compared the result with the unweighted estimate from all participants.

To account for equal sample sizes across zones of unequal population, we fitted each zone separately and combined estimates using population weights, treating zones as independent. Zone populations were summed from gridded population estimates across each zone’s health areas.^20^ For the overall rate, we combined zone-by-age-stratum estimates and accounted for within-household correlation between strata. Seropositivity was estimated with the same weights, with zones as strata and households as primary sampling units. Age-group weighting, heterogeneity testing, and sensitivity analyses using unpublished MSF programme populations or age-pooled kinetics are described in Supplementary Methods SM3.

We estimated exploratory HlyE IgA seroincidence by sex and self-reported typhoid history (restricted to ages ≥5 years), directly age-standardised because age distributions differed between subgroups. Subgroup estimates were compared using Wald tests on the log rate ratio with joint cluster-robust covariance (Supplementary Methods SM4). Zone and health-area rates, which the survey was not designed to compare, are reported in Supplementary Tables S2 and S3. We also modelled HlyE titres and IgA seropositivity as smooth functions of age using generalised additive models.

We estimated household-level HlyE IgA seropositivity using a random-intercept model with empirical-Bayes shrinkage and interpolated estimates by sample-size-weighted kernel smoothing with cross-validated bandwidth.

Post hoc, we estimated age-adjusted prevalence ratios for HlyE IgA seropositivity across recent illness groups (including malaria and/or anaemia) compared with no illness using the same models (Supplementary Methods SM5).

### Missing data

No values were imputed, and we retained the first record for 11 duplicate HlyE specimen identifiers. Subgroup analyses excluded 34 participants with missing sex and 3 participants aged ≥5 years with missing typhoid history (inclusion criteria in Supplementary Methods SM6).

### Ethical approval

The study protocol was approved by the MSF Ethics Review Board (ID 2216) and the Ethics Committee of the University of Kisangani, DRC (UNIKIS/CER/007/2022; amendments CER/005/GEAK/2023 and CER/002/GEAK/2024). Written informed consent was obtained from the head of each household and from each participating member, and children older than 10 years also gave assent. No names were collected; records were linked only to a household number and held on password-protected MSF servers in Europe (Supplementary Methods SM8).

## Results

### Participant enrolment and characteristics

Of 630 households targeted, 597 (94.8%) were surveyed: 22 were not reached and 11 declined; neither group was replaced (Figure 2). Of 2,814 participants with a recorded DBS-consent response, 2,768 (98.4%) consented. DBS was collected for 2,744 participants; 24 did not provide a DBS. Of the 2,744 DBS, 147 were not available for testing (134 not shipped, 13 not received by the laboratory), leaving 2,597 assay records. After removal of duplicate records, the seroincidence analysis included 2,586 participants in 582 households, of whom 2,504 (96.8%) were linked to a household location and included in the spatial analysis.

**Figure 2.**
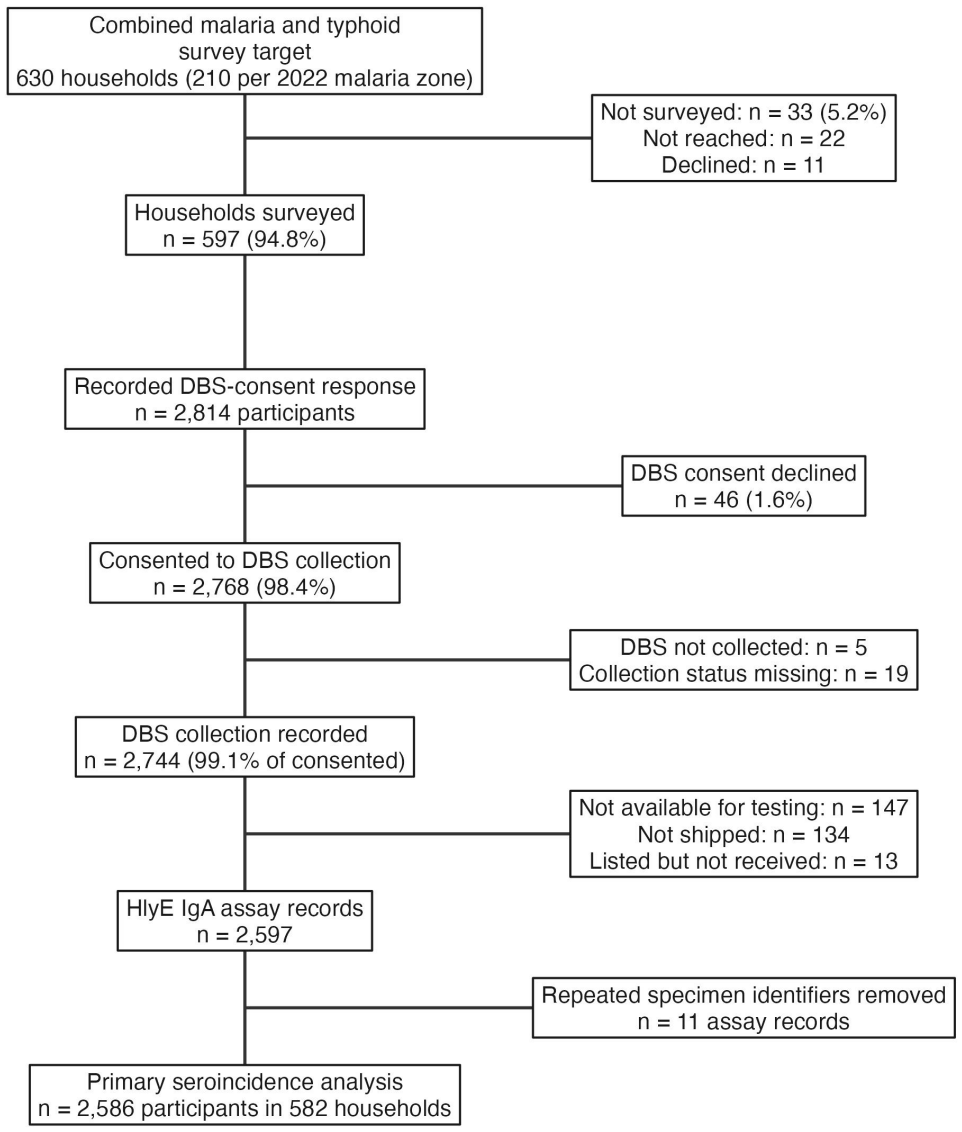
Participant flow, Angumu typhoid serosurvey, April–May 2024.

Of 2,586 participants, 1,106 (43%) were aged 16 years or older and 979 (38%) were aged 5– 15 years. Of the remainder, 371 (14%) were aged 2–4 years and 130 (5.0%) were younger than 2 years (Table 1; Supplementary Figure S2). Of participants with recorded sex, 1,393 (55%) were female. Prior typhoid fever was reported by 488 (19%) participants. Female participants were older than male participants (median age 15 vs 11 years). Participants reporting prior typhoid fever were older than those not reporting it (median age 22.5 vs 11 years; Supplementary Table S4).

### Primary HlyE IgA seroincidence

Overall HlyE IgA seroincidence, weighted by zone population, was 25 infections per 1,000 person-years (95% CI 22–29; Table 2). Seroincidence was highest among children younger than 2 years, at 91 infections per 1,000 person-years (95% CI 46–180). It was 22 (13–36) among children aged 2–4 years, 21 (17–26) at 5–15 years, and 28 (25–32) at 16 years or older. In an exploratory analysis by health area, seroincidence ranged from 20 to 46 infections per 1,000 person-years (Supplementary Table S3).

**Table 2.** Zone-weighted seroincidence of typhoidal *Salmonella* infection from HlyE IgA responses, by age group, sex and reported typhoid history.

| Group | Category | n | Seroincidence per 1,000 person-years (95% CI) | p value | Method |
|---|---|---|---|---|---|
| Age group | <2 | 130 | 91 (46–180) |  | Population-weighted |
| Age group | 2-4 | 371 | 22 (13–36) |  | Population-weighted |
| Age group | <5 | 501 | 29 (20–43) |  | Population-weighted |
| Age group | 5-15 | 979 | 21 (17–26) |  | Population-weighted |
| Age group | 16+ | 1106 | 28 (25–32) | <0.001 | Population-weighted |
| Sex | Female | 1393 | 22 (18–27) |  | Age-standardised |
| Sex | Male | 1159 | 28 (24–33) | 0.051 | Age-standardised |
| Self-reported typhoid history (age 5+) | No | 1616 | 24 (21–28) |  | Age-standardised |
| Self-reported typhoid history (age 5+) | Yes | 466 | 24 (19–31) | >0.99 | Age-standardised |
| Overall | Overall | 2586 | 25 (22–29) |  | Population-weighted |

In exploratory subgroup analyses, age-standardised HlyE IgA seroincidence was 22 infections per 1,000 person-years among female participants (95% CI 18–27) and 28 among male participants (24–33). The rate ratio was 0.80 (95% CI 0.64–1.00; p=0.051). Among participants aged 5 years or older, seroincidence was 24 per 1,000 person-years among those reporting a previous typhoid fever diagnosis (19–31). It was the same among those without such a history (21–28; p>0.99; Table 2).

### Spatial variation in HlyE IgA seropositivity

Weighted HlyE IgA seropositivity was 3.1% (95% CI 2.4–4.0), with 76 of 2,586 participants seropositive, all of whom were included in the spatial analysis. Smoothed seropositivity ranged from 2.4% to 7.2% across the study area (Figure 1B).

### Secondary and exploratory analyses

HlyE IgA seropositivity was 5.5% (25/454) among participants who reported an illness attributed to malaria and/or anaemia in the previous 2 weeks and had a negative malaria RDT at interview. It was 2.5% (37/1,454) among participants with no recent illness (age-adjusted prevalence ratio 1.98, 95% CI 1.21–3.25; p=0.006). The 25 seropositive participants in this group came from 24 different households. Seropositivity was 1.9% (6/320) among participants with the same illness and a positive RDT and 3.0% (8/266) among those with other illnesses (Supplementary Table S5).

The combined HlyE IgA and IgG model yielded an overall seroincidence estimate of 152 infections per 1,000 person-years (95% CI 146–159). By age group, the estimate was highest among children younger than 2 years (652, 95% CI 566–750) and lowest among participants aged 16 years or older (108, 95% CI 101–115; Supplementary Table S6).

HlyE IgG-only seroincidence was 399 infections per 1,000 person-years overall (95% CI 385–414) and exceeded both the IgA-only and combined-model estimates across all age groups (Supplementary Figure S3). HlyE IgA titres increased across the age range, whereas HlyE IgG titres increased until approximately 20 years of age and then remained stable.

Most IgA values were below the seropositivity cut-off, including in the oldest age group (Figure 3).

**Figure 3.**
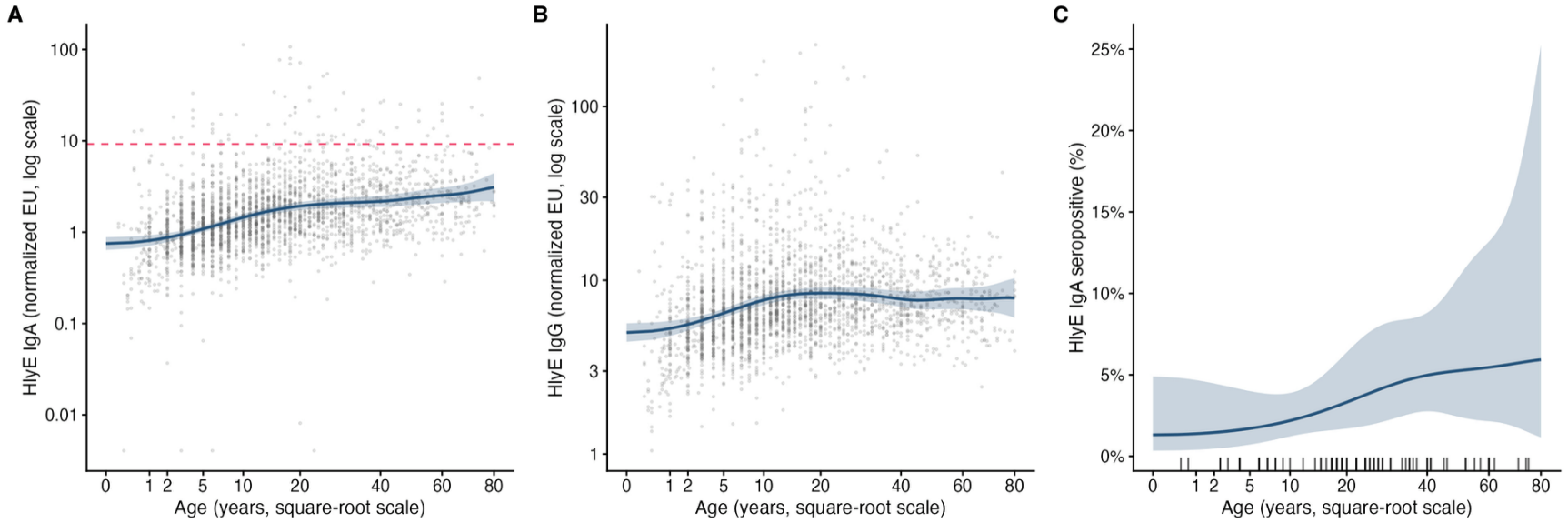
HlyE IgA and IgG titres and HlyE IgA seropositivity by age, Angumu Health Zone, DRC, April–May 2024. (A) HlyE IgA and (B) HlyE IgG titres in normalised ELISA units, log scale; points are individual participants. (C) HlyE IgA seropositivity; ticks mark seropositive participants. Lines, fitted values from generalised additive models; shaded areas, simultaneous 95% confidence bands. Dashed line, HlyE IgA seropositivity cut-off (Methods). Age is shown on a square-root scale.

In the one-participant-per-household sensitivity analysis, 582 participants were retained and the overall HlyE IgA seroincidence estimate was 26 infections per 1,000 person-years (95% CI 21–33; Supplementary Table S7), similar to the estimate of 28 (95% CI 25–31) from the unweighted analysis of all participants. With the MSF programme population figures in place of the WorldPop estimates, the weighted overall rate was 26 (95% CI 23–30). With each zone’s rate estimated using kinetics pooled across ages, the weighted overall rate was 28 (95% CI 25–32).

Alternative antibody-kinetics parameters changed the overall rate by factors of 0.92 to 1.83 relative to the reference, the largest change coming from kinetics derived from the Ghana cohort alone. Alternative measurement-noise assumptions changed it by less than 10% (Supplementary Table S8).

## Discussion

We report the first population-based seroincidence estimates of typhoidal *Salmonella* infection in the DRC. Among 2,586 participants, overall HlyE IgA seroincidence, weighted by zone population, was 25 per 1,000 person-years (95% CI 22–29). Seroincidence peaked among children younger than 2 years and remained above 20 per 1,000 person-years in every older age group. This pattern contrasts with SETA clinical surveillance in the DRC, where symptomatic typhoid peaked among school-aged children.^6^ This difference may reflect what each method measures: serology detects infection regardless of symptoms or care-seeking, whereas blood-culture surveillance detects only confirmed disease among patients investigated at health facilities.^26^ In the youngest children, infection may more often be subclinical or present as undifferentiated fever treated presumptively as malaria in this holoendemic setting. Consistent with this, HlyE IgA seropositivity was twice as high among participants with a recent illness attributed to malaria but a negative RDT as among those with no recent illness. The under-2 rate should also be interpreted with caution because few longitudinal antibody data exist for this age group. The kinetics applied came from 383 cases younger than 5 years, not resolved below 2 years.^4^ Among older children and adults, these rates are consistent with the 2022 Angumu verbal autopsy study. In that study, typhoid and paratyphoid accounted for 4.2% of deaths at ages 12 years and older.^18^

Comparisons with other published serosurveys require caution: populations, sampling designs, assay implementations, and model inputs differ sufficiently that direct ranking is of limited value. However, in a serosurvey in coastal and western Kenya,^27^ IgG responses to the same HlyE antigen were measured in the same laboratory and analysed with the same IgG-only model. Seroincidence in Angumu (399 per 1,000 person-years) was similar to that in coastal Kenya (370) and far higher than in western Kenya (36), where TCV has since been introduced.^27^ We re-estimated HlyE IgA seroincidence in Bangladesh, Nepal, and Pakistan from published data with the same model and age groups.^4^ At ages 5–15 years, Angumu seroincidence was higher than in Nepal (12 per 1,000 person-years) but lower than in Pakistan (85) and Bangladesh (260). The same order held in children younger than 5 years, whereas at ages 16 years or older, Angumu seroincidence was lower than in all three settings.

Estimates varied more than 14-fold by antibody choice. With kinetics pooled across ages throughout, so that the three are comparable, IgA alone yielded 28 per 1,000 person-years (95% CI 25–32), combined IgA/IgG 152 (146–159), and IgG alone 399 (385–414). HlyE IgA declines rapidly after infection and is more specific for recent infection.^4^ HlyE IgG persists longer and may accumulate with repeated infection, and its decay varies with local force of infection. IgG-based estimates are more sensitive to whether kinetics were derived from a population with comparable transmission intensity.^4^ We therefore consider IgA-based estimates the more conservative primary measure for Angumu. Raw IgG levels increased with age into early adulthood, as expected for an antibody that persists, but age-stratified models estimate recent-infection rates. Consequently, adding IgG raised estimates in every age group, and seroincidence remained highest among children younger than 2 years under each antibody choice.

Incorporating DBS-based typhoid serosurveillance into existing household surveys is operationally feasible^24^ and uses DBS collection, which is already routine in programmes such as HIV.^28^ However, HlyE serology and seroincidence models still require specialised laboratory and statistical capacity that currently limits routine use.

Seroincidence could be biased in either direction because no DRC antibody-kinetics data exist. IgA decay appears more comparable across settings,^22^ and alternative kinetic or noise inputs changed overall IgA seroincidence 0.92-to 1.83-fold (highest with Ghana kinetics). HlyE cannot distinguish *S*. Typhi from *S*. Paratyphi A (rarely isolated in sub-Saharan African enteric fever surveillance^4,6^), and Typhi-specific, TCV-preventable burden cannot be estimated. Seroincidence measures infection rather than clinical disease and cannot be mapped directly onto WHO clinical-incidence classifications.^11^ Because insecurity restricted sampling to 14 of 22 health areas, zone-wide seroincidence could be underestimated or overestimated if excluded areas had higher or lower transmission, respectively. CIs omit gridded-population uncertainty, although MSF programme weights yielded a similar rate (26 per 1,000 person-years, 23–30). Because sample size was calculated for the overall estimate only, age-specific precision was lower, lowest in the under-2 stratum (±74%). Underrepresentation of young adult men (Supplementary Figure S2) may underestimate seroincidence because eating outside the home increases risk.^29^ Broad age-band standardisation may leave residual confounding in sex- and typhoid-history estimates, with prior typhoid unconfirmed. With a median of four participants per household and no clustering test, the smoothed surface cannot confirm localised foci. However, a one-participant-per-household analysis yielded a similar unweighted rate (26 vs 28 per 1,000 person-years).

Protracted displacement, disrupted water and sanitation infrastructure, and the absence of bacteriological diagnostic capacity plausibly drive transmission in this setting. These conditions are widespread across Ituri and neighbouring provinces in eastern DRC. The main finding, that substantial typhoidal transmission can proceed largely undetected, is therefore likely to be generalisable across the region. Integrating DBS-based typhoid serosurveillance within existing operational household surveys, as done here, is operationally feasible and efficient. This approach generates evidence where dedicated surveillance infrastructure does not exist. The DRC has hosted a TCV mass vaccination campaign in Kisantu^30^ but has not introduced TCV into its national immunisation programme.^15^ Where direct incidence data are lacking, WHO’s 2026 position paper allows countries to infer probable typhoid incidence and severity from alternative data.^11^ Seroincidence estimates such as these could contribute to those data. These findings support broader, geographically representative, enteric fever surveillance across eastern DRC, linked where possible to culture or molecular confirmation, to shape the public health response. These seroincidence estimates show frequent infection in every age group. They also provide a baseline for evaluating any future TCV campaign in the health zone. Where surgical capacity is limited, preventing typhoid fever is likely to avert deaths from intestinal perforation. Clinicians at the hospital serving the health zone have observed this complication in large numbers.

## Acknowledgments

We thank the survey teams, community leaders, and participants in Angumu Health Zone.

## Contributors

JP, LG, IC, and EG designed the study and developed the protocol. JP, JS, and KA conducted the data analysis. LG, MM, PN, and BG coordinated field data collection. RC oversaw the laboratory analyses. All authors reviewed and approved the final manuscript.

## Registration

This observational study was not registered in a trial registry; participants were not assigned to any intervention by the investigators.

## Data sharing

Data used in this study are subject to restrictions arising from participant informed consent and privacy protections for identifying health information, and cannot be deposited in a public repository. De-identified individual-level data are available to qualified researchers on request to the Epicentre Data Sharing Committee (https://epicentre.msf.org/en/our-projects/study-data-access-request), which reviews requests independently of the study authors in accordance with Epicentre’s Data Sharing Policy (https://epicentre.msf.org/sites/default/files/2020-02/Epicentre-Data%20Sharing%20Policy-EN-2020.pdf). The study protocol is provided with this submission. Analysis code is available at https://github.com/jpolonsky/typhoid-angumu-2024, archived at Zenodo (https://doi.org/10.5281/zenodo.23762677).

