## Supplementary appendix for "Seroincidence of typhoidal *Salmonella* infection in eastern Democratic Republic of the Congo: a population-based cross-sectional serosurvey"

Contents: Supplementary Methods SM1–SM8; Supplementary Tables S1–S8; Supplementary Figures S1–S3. Reference numbers refer to the main text.

#### Supplementary Methods

##### SM1. Laboratory procedures

DBS were eluted into phosphate-buffered saline with 0.05% Tween overnight at 4 °C. DBS eluates were run in duplicate, with blank and positive-control wells in triplicate. Bound antibodies were detected using horseradish-peroxidase-conjugated goat anti-human IgA or IgG, with o-phenylenediamine substrate and kinetic absorbance readings at 450 nm. Blank-adjusted duplicate readings were averaged, divided by the human chimeric monoclonal HlyE antibody standard included on each plate, multiplied by 100, and reported as normalised ELISA units (EU).

##### SM2. Sampling frame

The sampling frame comprised the coordinates of all roofs in the accessible health areas, digitised from a February 2022 Airbus satellite image by Missing Maps and the MSF GIS Centre. Roofs were drawn at random from this frame and located in the field using GPS devices. For the host malaria survey, each drawn roof and its four nearest neighbours formed a cluster of five households. One household per cluster was enrolled in the typhoid serosurvey, giving 210 households per 2022 malaria intervention zone and 630 in total, with no allocation to health areas. In the specimens received, 582 households contributed at least one dried blood spot. Abandoned dwellings were not counted, and households that were absent at two visits or that declined to take part were not replaced.

##### SM3. Zone weighting

Each zone $z$ was fitted separately, giving a rate $\lambda_z$ with standard error $s_z$ on the log scale. The overall rate was $\lambda = \Sigma_z W_z \lambda_z$, with $W_z = N_z / \Sigma N$, where $N_z$ is the zone population. For an age group $a$, the weight of zone $z$ was proportional to $N_z n_{za} / n_z$, where $n_{za}$ is the number of participants of age group $a$ sampled in zone $z$ and $n_z$ the number sampled in the zone, normalised to sum to 1 within the age group. Zones were treated as independent and populations as known, so that $Var(\lambda) = \Sigma_z W_z^2 Var(\lambda_z)$, with $Var(\lambda_z)$ obtained by the delta method from $s_z$. The CI was calculated on the log scale.

In the main analysis, the overall rate was calculated from the nine zone-by-age-stratum estimates, each weighted by $(N_z / \Sigma N)(n_{za} / n_z)$. Its variance used the joint covariance of these estimates, because strata within a zone include members of the same households. The alternative overall estimate was calculated from the three zone estimates fitted with the kinetics pooled across ages. The two estimates differ because the pooled and age-specific kinetics are different parameter sets, not because of the weights. The ratio of the alternative to the main estimate was similar within each zone (1.123, 1.114, and 1.115) and in the unweighted estimates (1.116). Overall rates that included HlyE IgG were calculated from the three zone estimates fitted with the kinetics pooled across ages, because the joint covariance of their age-stratum estimates was not available.

Zone populations were the sums of the WorldPop/GRID3 bottom-up gridded population estimates, version 4.4 (approximately 100 m resolution), over the polygons of each zone's health areas, weighting each grid cell by the fraction of its area inside the polygon (reference 20).

Before analysis, we specified that a zone-by-age estimate would be considered unstable if the optimiser did not converge cleanly, the Hessian was not positive, or the standard error of the log rate exceeded 2. An unstable estimate for the group under 2 or 2 to 4 years would then be replaced by the combined estimate for children under 5. Unweighted estimates from a single fit of all participants are reported as a check.

Age-group rates were combined across zones in the same way, and heterogeneity across the four age groups was tested with Cochran's Q statistic on the log rates, treating age groups as independent. In sensitivity analyses, we recalculated the weighted overall rate with unpublished MSF programme population estimates in place of the gridded estimates, and from zone-specific fits with kinetics pooled across ages.

#### SM4. Age standardisation

The sex and typhoid-history comparisons were age-standardised if the share of any of the four age groups differed by 5 percentage points or more between the groups compared, or if a chi-square test of the age-group distributions gave $p<0.05$. We directly age-standardised the subgroup rates to the zone-weighted age distribution of all participants, because the age distribution differed between the sexes and between participants with and without a reported history of typhoid fever. Rates by sex and by history were estimated within each age stratum, pooled across zones, with household-clustered standard errors. The stratum rates were combined with the weights of the standard population, renormalised over the strata 5 to 15 and 16 years or older for history. Variances used the joint cluster-robust covariance of all the stratum estimates of a comparison, since participants of different groups and strata share households. Crude rates from the kinetics pooled across ages are reported as a check.

#### SM5. Recent illness group definitions

We compared HlyE IgA seropositivity between participants who reported an illness they attributed to malaria and/or anaemia in the 2 weeks before the interview and tested negative on a malaria RDT at the interview, and participants with no illness in that period.

Participants with the same illness and a positive RDT, and those with any other illness, were compared with the same reference group.

#### SM6. Analysis inclusion criteria

Primary seroincidence analyses included participants with an HlyE IgA result and recorded age. Spatial analyses were restricted to participants linked to a household location.

#### SM7. Age ascertainment

Age was based on self-report; for young children, a parent or guardian reported age on their behalf.

#### SM8. Data protection

No participant names were collected. Individual records were linked only to a household number. Data were encrypted on the collection tablets and transferred directly to an MSF-hosted KoBoToolbox server in Europe, with subsequent storage on MSF-licensed servers in Europe. Access was limited to the study investigators and the medical coordinator, and the data will be destroyed five years after collection. Dried blood spots were destroyed after testing. Household coordinates are not released, and published maps show no household locations.

### Supplementary Tables

**Supplementary Table S1. Antibody kinetic and measurement-noise parameters.** (A) Seroresponse kinetics, median (2.5th–97.5th percentile) of 40,000 posterior draws, pooled from the Bangladesh, Ghana, Nepal, and Pakistan cohorts (age groups with no participants from a cohort draw on the others). y0, baseline titre; y1, peak titre (ELISA units); t1, time to peak (days); alpha, decay rate (per day); r, decay shape. The overall parameters were used for estimates pooled across ages.

| Antibody | Age group, years | y0 | y1 | t1 | alpha | r |
|---|---|---|---|---|---|---|
| HlyE IgA | <5 | 20.8 (2.43-181) | 37.6 (6.95-470) | 28.5 (8.01-85.1) | 9.03e-05 (1.82e-06-0.00398) | 3.24 (1.84-6.95) |
| HlyE IgA | 5–15 | 14.9 (0.712-295) | 103 (13-1,390) | 20 (6.4-54) | 9.41e-05 (1.29e-06-0.0062) | 2.58 (1.7-4.54) |
| HlyE IgA | ≥16 | 14 (0.663-290) | 131 (10.1-2,570) | 20.8 (5.99-66.4) | 6.21e-05 (2.31e-06-0.0014) | 2.64 (1.86-4.13) |

| Antibody | Age group, years | y0 | y1 | t1 | alpha | r |
|---|---|---|---|---|---|---|
| HlyE IgA | Overall | 13.8 (0.753-255) | 94 (14.1-892) | 19.6 (5.15-70.8) | 6.91e-05 (9.52e-07-0.00498) | 2.81 (1.69-5.69) |
| HlyE IgG | <5 | 14.7 (1.21-181) | 160 (20.7-1,690) | 16.2 (2.27-105) | 0.00011 (5.08e-06-0.00168) | 2.2 (1.63-3.22) |
| HlyE IgG | 5–15 | 15.1 (0.72-313) | 223 (49.6-1,540) | 12.3 (3.5-41.4) | 7.59e-05 (2.31e-06-0.00213) | 2.08 (1.38-4) |
| HlyE IgG | ≥16 | 15.6 (0.659-381) | 249 (30-2,730) | 19.1 (6.44-46.8) | 1e-04 (9.97e-06-0.000954) | 2.01 (1.4-3.4) |
| HlyE IgG | Overall | 14.5 (0.628-324) | 226 (56.5-1,100) | 15.6 (3.75-63.7) | 6.3e-05 (2.33e-06-0.00161) | 2.18 (1.48-3.88) |

(B) Measurement-noise parameters by laboratory. nu, biological noise; eps, measurement noise; y.low and y.high, lower and upper limits of detection (ELISA units). The parameters of the laboratory that tested the Angumu samples were used; the others inform the sensitivity analysis in Supplementary Table S8.

| Antibody | Laboratory | nu | eps | y.low | y.high |
|---|---|---|---|---|---|
| HlyE IgA | CHRF (Bangladesh) | 2.6 | 0.28 | 0.376 | 1,000 |
| HlyE IgA | MGH (Ghana) (used) | 2.6 | 0.24 | 0.179 | 1,000 |
| HlyE IgA | DH (Nepal) | 2.6 | 0.238 | 0.853 | 1,000 |
| HlyE IgA | AKU (Pakistan) | 2.6 | 0.279 | 0.508 | 1,000 |
| HlyE IgG | CHRF (Bangladesh) | 2.36 | 0.306 | 0.787 | 1,000 |
| HlyE IgG | MGH (Ghana) (used) | 2.36 | 0.164 | 0.645 | 1,000 |
| HlyE IgG | DH (Nepal) | 2.36 | 0.128 | 1.89 | 1,000 |
| HlyE IgG | AKU (Pakistan) | 2.36 | 0.146 | 1.59 | 1,000 |

**Supplementary Table S2. HlyE IgA seroincidence by 2022 malaria intervention zone and age group.** Infections per 1,000 person-years (95% CI), household-clustered standard errors. Zone population, WorldPop/GRID3 version 4.4 gridded estimate summed over the zone's sampled health areas. Zone overall rates use kinetics pooled across ages. The survey was not designed to compare zones.

| Zone | Age group, years | n | Seroincidence (95% CI) | Zone population |
|---|---|---|---|---|
| Routine | <2 | 39 | 86 (27–273) | |

| Zone | Age group, years | n | Seroincidence (95% CI) | Zone population |
|---|---|---|---|---|
| | 2–4 | 120 | 19 (7–50) | |
| | 5–15 | 352 | 15 (10–22) | |
| | ≥16 | 368 | 25 (20–31) | |
| | Overall | 879 | 24 (20–29) | 76,760 |
| IRS | <2 | 50 | 50 (12–200) | |
| | 2–4 | 129 | 20 (9–43) | |
| | 5–15 | 328 | 20 (14–29) | |
| | ≥16 | 377 | 23 (18–29) | |
| | Overall | 884 | 25 (20–30) | 35,152 |
| MDA+IRS | <2 | 41 | 123 (45–337) | |
| | 2–4 | 122 | 26 (13–53) | |
| | 5–15 | 299 | 29 (21–41) | |
| | ≥16 | 361 | 35 (28–43) | |
| | Overall | 823 | 37 (31–44) | 59,880 |

**Supplementary Table S3. Exploratory HlyE IgA seroincidence by health area.** Infections per 1,000 person-years (95% CI), kinetics pooled across ages, household-clustered standard errors. Unweighted. The survey was not designed to compare health areas.

| Zone | Health area | Households | Participants | IgA seropositive | Seroincidence (95% CI) |
|---|---|---|---|---|---|
| Routine | Abia | 45 | 191 | 3 | 30 (20–45) |
| Routine | Ara | 28 | 104 | 2 | 21 (13–34) |
| Routine | Awasi | 51 | 267 | 6 | 20 (15–27) |
| Routine | Cawa | 52 | 204 | 9 | 26 (17–40) |
| Routine | Linga | 23 | 113 | 5 | 21 (11–40) |
| IRS | Anzika | 41 | 175 | 3 | 29 (19–45) |
| IRS | Pakala | 55 | 279 | 5 | 23 (17–32) |
| IRS | Panyadong | 38 | 210 | 2 | 23 (17–32) |
| IRS | Udongo Abira | 45 | 198 | 7 | 26 (16–41) |
| MDA+IRS | Apala | 32 | 146 | 8 | 45 (29–69) |
| MDA+IRS | Gengere | 46 | 174 | 2 | 26 (17–39) |
| MDA+IRS | Lanyi | 27 | 124 | 4 | 31 (18–55) |
| MDA+IRS | Ugudo-Zii | 52 | 235 | 11 | 46 (32–65) |

| Zone | Health area | Households | Participants | IgA seropositive | Seroincidence (95% CI) |
|---|---|---|---|---|---|
| MDA+IRS | Uyandu | 33 | 133 | 9 | 36 (25–54) |

**Supplementary Table S4. Age distribution by sex and by self-reported typhoid history.** Participants with recorded sex, and with a recorded typhoid history, respectively. Both comparisons met the age-standardisation criterion (Supplementary Methods SM4).

| | Female | Male | No typhoid history | Typhoid history |
|---|---|---|---|---|
| Participants, n | 1,393 | 1,159 | 2,093 | 488 |
| <2 years, n (%) | 69 (5%) | 54 (4.7%) | 127 (6.1%) | 3 (0.6%) |
| 2–4 years, n (%) | 174 (12.5%) | 194 (16.7%) | 350 (16.7%) | 19 (3.9%) |
| 5–15 years, n (%) | 469 (33.7%) | 487 (42%) | 842 (40.2%) | 136 (27.9%) |
| ≥16 years, n (%) | 681 (48.9%) | 424 (36.6%) | 774 (37%) | 330 (67.6%) |
| Median (IQR) age, years | 15 (6–29) | 11 (5–22) | 11 (5–23) | 22.5 (13–37) |
| Chi-square p, age-group distribution | <0.001 | | <0.001 | |
| Largest difference in age-group share, percentage points | 12.3 | | 30.6 | |

**Supplementary Table S5. HlyE IgA seropositivity by illness in the 2 weeks before the interview.** Age-group-adjusted prevalence ratios from log-binomial regression with household-clustered robust standard errors (Supplementary Methods SM5). Malaria and/or anaemia, illness attributed by the respondent to malaria and/or anaemia; RDT, malaria rapid diagnostic test at the interview.

| Group | Participants | IgA seropositive, n (%) | Prevalence ratio (95% CI) | p |
|---|---|---|---|---|
| No illness in past 2 weeks | 1,454 | 37 (2.5%) | 1 (reference) | |
| Malaria/anaemia, RDT-negative | 454 | 25 (5.5%) | 1.98 (1.21–3.25) | 0.0063 |
| Malaria/anaemia, RDT-positive | 320 | 6 (1.9%) | 0.85 (0.36–1.98) | 0.71 |
| Other illness | 266 | 8 (3.0%) | 1.12 (0.54–2.33) | 0.75 |

**Supplementary Table S6. Combined HlyE IgA and IgG seroincidence by age group.** Infections per 1,000 person-years (95% CI), household-clustered standard errors. Zone-weighted rates combine per-zone fits with kinetics pooled across ages (Supplementary Methods SM3).

| Age group, years | Zone-weighted | Unweighted |
|---|---|---|
| <2 | 652 (566–750) | 643 (566–731) |
| 2–4 | 272 (257–288) | 273 (259–288) |
| <5 | 318 (299–338) | 319 (301–337) |
| 5–15 | 137 (131–143) | 138 (132–143) |
| ≥16 | 108 (101–115) | 108 (102–115) |
| Overall | 152 (146–159) | 153 (147–159) |

**Supplementary Table S7. HlyE IgA seroincidence with one participant per household.** One participant drawn at random from each of the 582 households; unweighted, infections per 1,000 person-years (95% CI).

| Age group, years | Participants | Seroincidence (95% CI) |
|---|---|---|
| <2 | 31 | 177 (66–475) |
| 2–4 | 80 | 7 (1–44) |
| 5–15 | 198 | 20 (13–31) |
| ≥16 | 273 | 25 (19–32) |
| Overall | 582 | 26 (21–33) |

**Supplementary Table S8. Sensitivity of HlyE IgA seroincidence to the antibody-kinetic and measurement-noise inputs.** Unweighted analysis of all 2,586 participants, household-clustered standard errors. Infections per 1,000 person-years, with the ratio to the reference (K0) in parentheses. Ratios apply approximately to the zone-weighted estimates.

| Scenario | Description | <2 | 2–4 | 5–15 | ≥16 | Overall |
|---|---|---|---|---|---|---|
| K0 | Reference: pooled kinetics (all cohorts), testing-laboratory noise | 84 | 22 | 21 | 27 | 27 |
| K1 | Ghana-only kinetics | 88 (1.06) | 28 (1.30) | 38 (1.82) | 62 (2.25) | 49 (1.83) |
| K2 | Pakistan-only kinetics | 84 (1.00) | 22 (1.04) | 19 (0.93) | 23 (0.85) | 25 (0.92) |
| K3 | OSF rtw5k kinetics, not age-stratified | 82 (0.98) | 21 (0.96) | 26 (1.26) | 58 (2.12) | 42 (1.56) |
| K4 | 5-15-year non-hospitalised | 82 | 20 | 22 | 40 | 32 |

| Scen ario | Description | <2 | 2–4 | 5–15 | ≥16 | Overall |
|---|---|---|---|---|---|---|
| | kinetics, all ages | (0.98) | (0.93) | (1.06) | (1.46) | (1.20) |
| K5 | 5-15-year culture-confirmed *S.* Typhi kinetics, all ages | 83 (0.99) | 20 (0.94) | 21 (1.02) | 35 (1.28) | 30 (1.12) |
| N1 | Worst laboratory noise (eps and y.low maxima across the 4 laboratories) | 84 (1.00) | 20 (0.92) | 20 (0.95) | 26 (0.96) | 26 (0.96) |
| N2 | Testing-laboratory measurement noise (eps) doubled | 77 (0.92) | 16 (0.72) | 18 (0.86) | 29 (1.05) | 25 (0.93) |

### Supplementary Figures

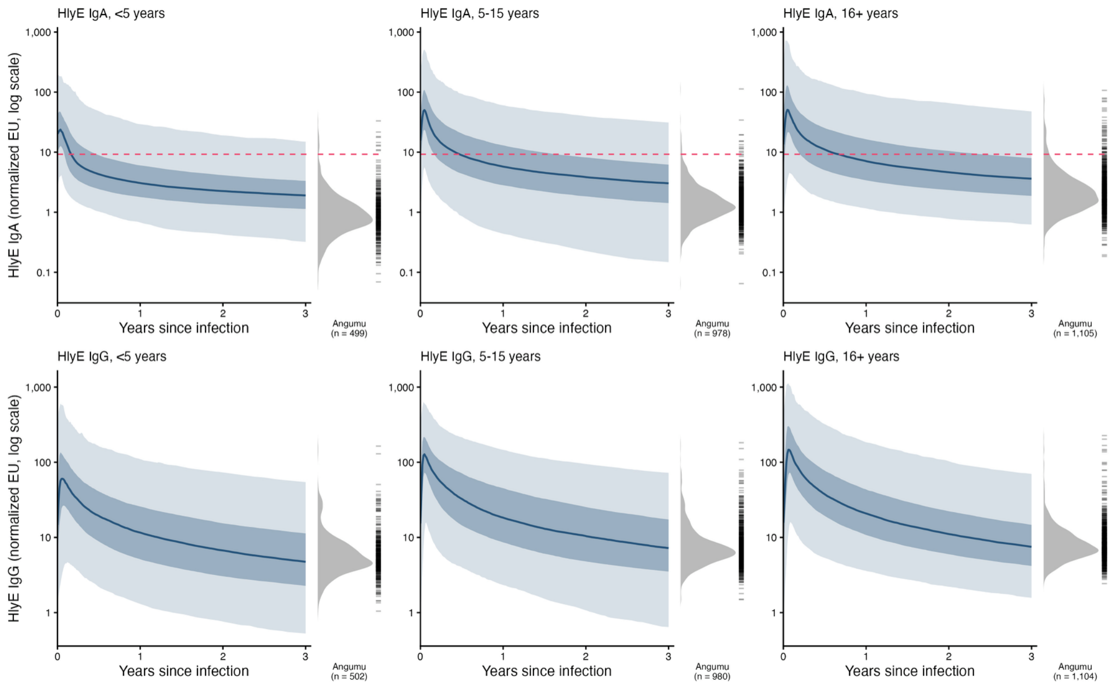


**Supplementary Figure S1. Assumed HlyE IgA and IgG antibody responses after infection, with the Angumu titres.** Lines, median of the posterior kinetics by age group; dark and light bands, 50% and 95% intervals. Right of each panel, density and individual values of the Angumu titres for that age group (titres of zero, 4 IgA values, cannot be shown on the log scale). Dashed line, HlyE IgA seropositivity cut-off.

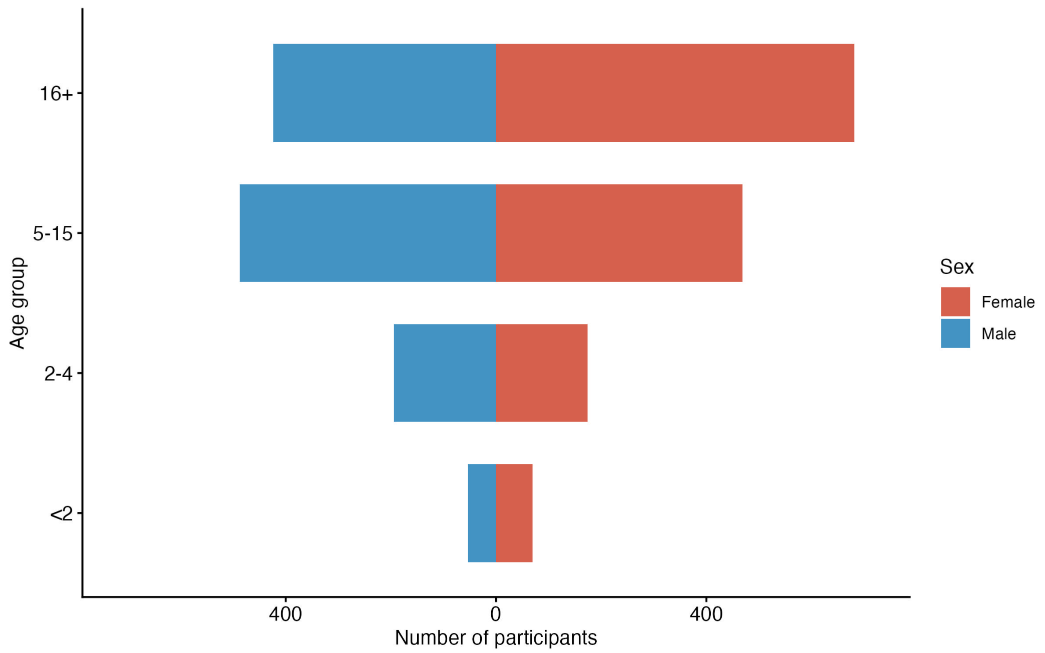


**Supplementary Figure S2. Participants by age group and sex.** 2,552 participants with recorded sex.

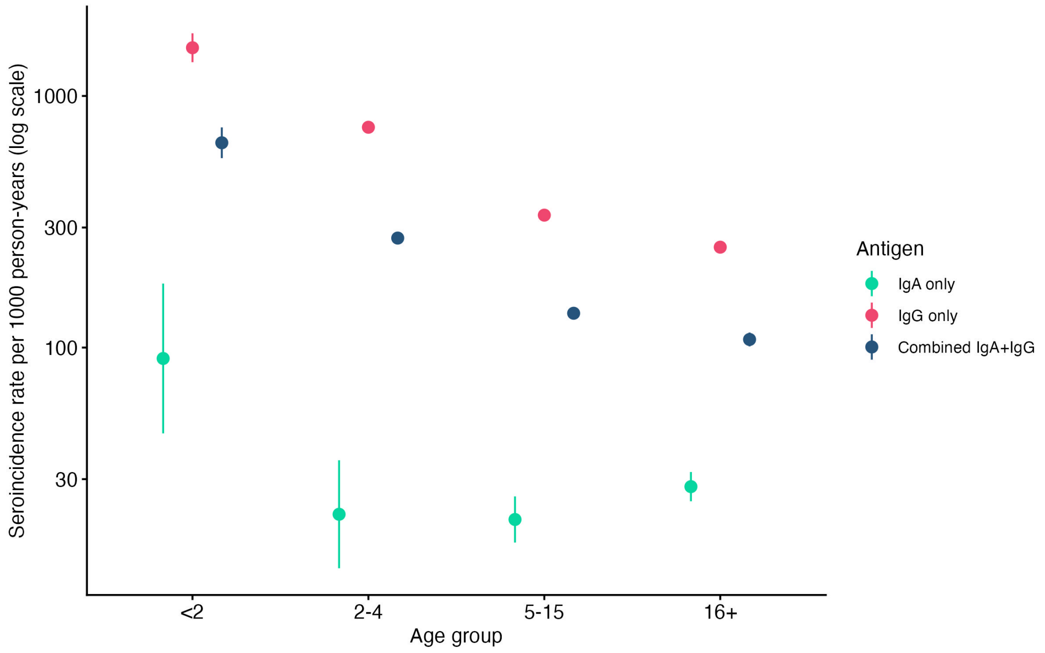


**Supplementary Figure S3. Zone-weighted seroincidence by age group, estimated from HlyE IgA only, IgG only, and both combined.** Points, estimates; bars, 95% CIs.
